# A methodology for selection and quality control of the radiological computer vision deployment at the megalopolis scale

**DOI:** 10.1101/2022.02.12.22270663

**Authors:** Anna E. Andreychenko, Tatiana A. Logunova, Victor A. Gombolevskiy, Aleksandr E. Nikolaev, Anton V. Vladzymyrskyy, Valentin E. Sinitsyn, Sergey P. Morozov

**Affiliations:** Research and Practical Clinical Center for Diagnostics and Telemedicine Technologies of the Moscow Healthcare Department

**Keywords:** Radiology, Artificial Intelligence, Quality Control, Health Plan, Implementation Innovation, Health Knowledge, Attitudes, Practice

## Abstract

In recent years, there has been tremendous interest in the use of artificial intelligence (AI) in radiology in order to automate the interpretation. However, uncontrolled and widespread use of AI solutions may have negative consequences. Therefore, before implementing such technologies in healthcare, thorough training of personnel, adaptation of information systems, and standardized datasets for an external validation are required. All this necessitates a formation of a unique unified methodology. The best practices of AI introduction in diagnostic radiology are still subject to debate and require new results of a scientific-practical research with the assessment of implementation conditions.

This work discusses expected issues and potential solutions for the introduction of computer vision-based technologies for automatic analysis of radiological examinations with an emphasis on the real-life experience gained during simultaneous AI implementation into practice of more than a hundred state radiology departments in 2020-2021 in Moscow, Russia (an experiment). The experiment used end-user software testing approaches, quality assurance of AI-based radiological solutions, and accuracy assessment of the AI-empowered diagnostic tools on local data. The methods were adapted and optimized to ensure a successful real-life radiological AI deployment on the extraordinary large scale. The experiment involved in total around thousand diagnostic devices and thousand radiologists. AI deployment was associated with additional options in a routine radiologist’s workflow: triage; additional series formed by AI with indication of pathological findings and their classification; report template prepared by AI in accordance with the target clinical task, user feedback on AI performance.

A multi-stage methodology for implementing AI into radiological practice that was developed and advanced during the experiment is described in this report.

**Essentials:**

- A methodology for the AI deployment for non-academic radiological sites excluded more than half of the offered AI solutions that do not fulfill the diagnostic and functional requirements
- Quality control of AI should be supported by not only data scientists, IT specialists or engineers, but also by radiologists at all stages of selection and testing.
- Radiologists need to understand the capabilities, limitations of AI by getting an additional training.

## Introduction

Introduction of artificial intelligence(AI) into the diagnostic radiology is primarily aimed at automating interpretation and analysis of medical images. Diagnostic radiology differs from other medical specialties by a high digitalization with many opportunities for the machine learning developments. A joint statement by the International Society of Radiographers and Radiological Technologists(ISRRT) and the European Federation of Radiographer Societies(EFRS) states that AI can optimize imaging workflows and potentially help reduce a radiation dose, improve research efficacy and consistently ensure high-quality planning processes, gradually turning AI into the integral working tool(1-4). Multiple publications of American College of Radiology and the British Institute of Radiology describe the possible benefits of AI implementation in a routine radiological practice(5,6). In a 2019, an AI-dependent decrease in the radiologists’ workload was noted only in 5% of publications; in the rest of the assessments, radiologist efforts increased due to the need to learn a new software and increase in the reporting time due to reading the additional results from AI(7). The same time the effective implementation of AI in medical facilities is associated with technical difficulties and reliable encryption of the received data when transmitting them outside the clinic(7), as well as lack of funds, regulatory policies, and support systems(8). On the other hand, a cornerstone of the AI implementation should be scientific and clinical validity, relevance to the intended purpose, and user-friendliness, which can reduce labor costs of the radiologists and increase the efficacy of radiological reports, i.e. achieve the initial goal of automating processes with application of AI.

These issues were discussed in the statement of North American and European radiologists(9) and were formulated by Recht and co-authors in the consolidated methodological recommendations that address the ethical aspect in detail, however, mention only general technical requirements for the integration process of AI into diagnostic radiology(10). In the absence of the detailed practical recommendations for a radiological AI deployment, we have developed and tested a systematic methodology for integration of AI into Moscow Unified Radiological Information Service(URIS), which unites all non-academic state medical radiological facilities of the megalopolis.

### Methodology description

There are hundreds of AI solutions for diagnostic imaging available on the market. On the one hand, the variety of software is an advantage for users in choosing the most suitable option. On the other hand, there are no regulated principles for choosing a right solution for various tasks, considering specifics of the medical facility and healthcare system, as well as variable environmental conditions (e.g., the epidemiological situation)(11).

In the Moscow healthcare system in 2020, the experiment was launched to introduce innovative computer vision technologies into a practice of radiology departments for analysis of medical images. The study is registered on clinicaltrials.gov with the number NCT04489992 with the Center for Diagnostics and Telemedicine Technologies as the experiment’s organizer. A purpose of the Experiment was a scientific and practical study of the possibility of using decision-making support methods based on data analysis results with the application of advanced innovative technologies in the Moscow healthcare system. The experiment was based on the methodology of software testing by end users – radiologists(12). URIS became the infrastructural backbone for the experiment(13) that unites more than 1000 radiologists with 192 CT scanners, 119 mammography units, 630 X-ray units of the state radiological departments.

Three most common routine modalities in URIS were selected for the AI deployment: chest CT, chest X-ray, and mammography. The experiment was widely communicated in the professional community RSNA 2019 and personalized invitations were sent out to AI radiology companies to participate. The list of company invitations was obtained from Internet searches for the keywords “AI”, “artificial intelligence”, “machine learning”, “radiology”, “x-ray”, “mammography” and “CT”. Inclusion criteria were: developer’s statement about the radiological use case, contact information. Exclusion criteria were: scientific or non-commercial organization, no response to the invitation within 1 month, absence of own computing facilities for the data processing, legal restrictions on work in Russia. For invited developer, the AI functionality and diagnostic accuracy metrics were tested. Several local datasets were used to standardize the experiment in the best possible way. Based on testing results, we collected feedback from the user-radiologists, obtained by integrating feedback forms into URIS for each AI solution. All AI companies received grants from the state after 3 months of prospective implementation for each study analyzed. Also, an expert audit of sample studies was conducted.

Currently, there are no universal standards for the implementation of AI-based products in the routine practice of medical facilities of the megalopolis. Unlike studies based on data of academic medical centers, when AI was primarily tested by experts on a limited number of specific exams, the proposed methodology was applied for the first time in the city healthcare system with 252 inpatient and outpatient radiology departments equipped with 295 digital diagnostic devices compatible with AI solutions. The enormous volume and heterogeneity of data, broad-functionality of clinics and wide variations in the skill level of radiologists required a thorough step-by-step assessment and rigorous selection of the proposed AI products, as well as periodic quality control, which was carried out by both data scientists and radiologists from a practical end-user point of view(4). The methodology of the experiment is structured in **Table 1**, describing the stages and involved participants. In total 40 AIs provided by 21 developers took part in the experiment during 2020; 18 AI models were integrated into URIS with following assessment. AI models analyzed 1468872 studies: chest CT – 56,0%(818296), mammography – 4,0%(61497), chest X-ray – 18,0%(270965), chest fluorography – 22,0%(318114).

**Table 1.** Proposed methodology stages for the selection and assessment of AI solutions for diagnostic radiology in the process of their introduction into a routine practice.

|  | Stage | Means | Participants | Purpose | Results |
| --- | --- | --- | --- | --- | --- |
| **0** | Basic diagnostic task | Consensus of radiologists on necessary features for the use case | Engineers (data scientists) and radiologists | Define standard output that AI should provide | Unified requirements for AI |
| **1** | Self-test | with DICOM files examples of the devices models | AI developer | check technical compatibility of an AI to analyze the DICOM files correctly | Decision on integration |
| **2** | Functional test | 5 selected studies from PACS | Engineers (data scientists) and radiologists | evaluate the AI service from the user's point of view (radiologist); ensure correct integration | seamless integration, convenience, and ease of use availability and accessibility of all elements of AI results, patient safety. |
| **3** | Calibration test | 100 selected studies from PACS | Engineers (data scientists) | Evaluate claimed accuracy metrics on the local data, optimize threshold | Confirmed diagnostic accuracy.<br>Threshold<br>Processing |
|  |  |  |  | value,<br>AI analysis<br>duration | time |
| **4** | QC &<br>Monitoring | Selected<br>studies,<br>analyzed by<br>AI real-time | Engineers (data<br>scientists) and<br>radiologists | Ensure proper<br>and stable<br>performance<br>on each<br>diagnostic<br>device | Real-world<br>technical and<br>diagnostic<br>defects<br>detection and<br>correction |
| **5** | Radiologists<br>feedback &<br>expert audit | Feedback<br>form in<br>PACS;<br>check-list,<br>expert audit | Radiologists &<br>experts | Assess<br>practical and<br>diagnostic<br>value of<br>AI | End users and<br>expert<br>evaluation |

### Stages of the AI model selection

#### 0. A preparatory stage

aimed to define clearly a baseline diagnostic task for an AI model per use case included in the experiment. Firstly, requests were sent to companies that provided AI solutions in radiology, to specify diagnostic task’s details of the solutions. Taking into account the received answers experienced (more than 10 years) radiologists formulated baseline diagnostic tasks(**Table 2**), for which radiologists can potentially benefit from AI in terms of the report turn-around time reduction and increase of the diagnostic accuracy. With the support of AI developers, the minimum required functionality for the AI solutions and the convenient report form were determined. As a result, general requirements for the AI functionality were formed(**Table 3**). Involvement of the end user in the development of baseline requirements at the preparatory stage ensured exclusion of the AI solutions that would obviously not be in demand and convenient in practice.

**Table 2.** A list of use cases selected by the radiologist as the most promising ones for AI deployment at the non-academic radiological departments.

| № | Modality and anatomy | Diagnostic goal |
| --- | --- | --- |
| 1 | Chest CT | Detection of coronavirus infection (COVID-19) signs |
| 2 | Chest CT/LDCT | Detection of lung cancer signs (ICD-10 code C34) |
| 3 | Chest X-ray and fluorography | Detection and localization of signs correlating with at least one of the priority nosologies according to the ICD-10 classification: A15-A16, A19, C34-C39, D38.1-D38.4, J10-J18, J80-J86, J98.1, J94, R09.1, S22, S27.0 |
| 4 | Screening/diagnostic mammography | Detection and localization of breast cancer signs (ICD-10 code C50) |

**Table 3.** Baseline requirements for AI’s functionality in RIS

|  |  |
| --- | --- |
| Worklist | Triage is a prioritization of the analysis and report of studies in which AI identified changes |
|  | Labeling of AI-processed studies |
| Image visualization | Creation of the additional series of images with AI results. The original images should be unchanged.<br>*Alternative: fully switchable overlay with labeling of findings.* |
|  | Recognizable name of the additional series with AI results.<br>*Alternative: if the results are in overlay format, the information about it should be added to the series name.* |
|  | The images should have the following information:<br>- name and version of AI software,<br>- probability of the target pathology,<br>- warning "For research purposes only". |
| AI's structured report | 1. Purpose of AI solution (*corresponds to the diagnostic task*) |
|  | 2. General information about the AI model, developer, date and time of the analysis |
|  | 3. Quick user guide |
|  | 4. Conclusion and details of the analysis |

#### 1. AI self-testing

To check technical compatibility with the DICOM files the AI solutions providers that fulfilled the baseline and diagnostic and functional requirements were offered a small dataset with samples from all type of diagnostic devices per use cases that were in the URIS. The self-testing was carried out on a provider’s side without control from the experiment’s organizer. Compatibility of the AI and the self-test dataset allowed to proceed to further stages of AI evaluation and its integration into the testing environment of URIS (“URIS-test”) “URIS-test” was a replica of the URIS but was isolated from the real-world data.

The further steps of AI assessment were carried out on a side of the experiment’s organizer. The functional and calibration testing were carried out in the “URIS-test”. At this stage, there was no direct access to patient data, but to prepared anonymized datasets imported from URIS. If an AI solution passed this stage successfully, it was then integrated to URIS. Technical testing, monitoring, feedback collection and clinical audit were carried out during the real-life AI validation in URIS.

#### 2. Functionality testing

Testing was carried out on a dataset of 5 studies anonymized and imported from the URIS. This dataset included 2 studies with target pathology, 2 studies without target pathology according to the baseline diagnostic task, and 1 with a defect (e.g., the lungs were not fully scanned). The presence and absence of the radiological signs of the target pathologies in the studies were confirmed by consensus of 5 certified radiologists with more than 5 years of experience. The testing was aimed to check the integration and functionality completeness, convenience and ease of use by a radiologist of all of the AI generated elements in the URIS worklist, image viewer and reporting area. In 2020, 40 AI were tested.

The tested functionality included sorting of the worklist (triage), highlighting pathological findings on the images, and filling out a structured report.

For **AI-based management of a worklist**, two functions were necessary:

– execution of the triage – prioritization and labeling of studies with pathological findings revealed by AI;
– presence of unambiguous labeling in the worklist, that allows a radiologist to understand whether a study was already processed by AI and whether pathological changes were detected.

*RIS of different manufacturers may have a manual prioritization function at different levels – by a referring physician, by a radiographer, by a radiologist. If the prioritization was done manually, the AI triage function may interfere and have unwanted negative consequences. IT-specialists should account for elimination of such possible negative interference during the AI integration into RIS*.

Mandatory AI functionality in the PACS viewer **when working with images** included:

– availability of a separate series with the images processed by the AI;
– clear and unambiguous labeling of pathological findings in the image (heat map, color contouring, contrast contouring, etc. – labeling method was at the discretion of the developer);
– indicating information about AI (name and a software version) on the images processed by the AI, a probability of each finding, and the warning “For research purposes only”;
– in the absence of a separate series, labeling could be applied to the original series with the indication of this in the name of the series, while the original data (date and time, patient information, etc.) were always preserved on the image and there was a possibility of disabling the overlay.

Image visualization could be incorrect due to distortion during processing by AI and/or incomplete delivery of the study. Some information, such as colored labeling, can be lost when using monochrome diagnostic monitors (for example, in a mammography room). AI developer is responsible for the elimination of these inconsistencies.

*Involvement of radiologists at this stage allowed recognition of the inconsistencies that might be missed by IT-specialists and engineers, e.g. when findings are located outside the target anatomical region **(Fig.1a, b, c)**, or the detected changes do not relate to the diagnostic task **(Fig. 1d)**. IT-specialists/engineers are able to recognize the obvious technical defects caused by AI malfunctioning **(Fig. 2)**.*

**Figure 1.**
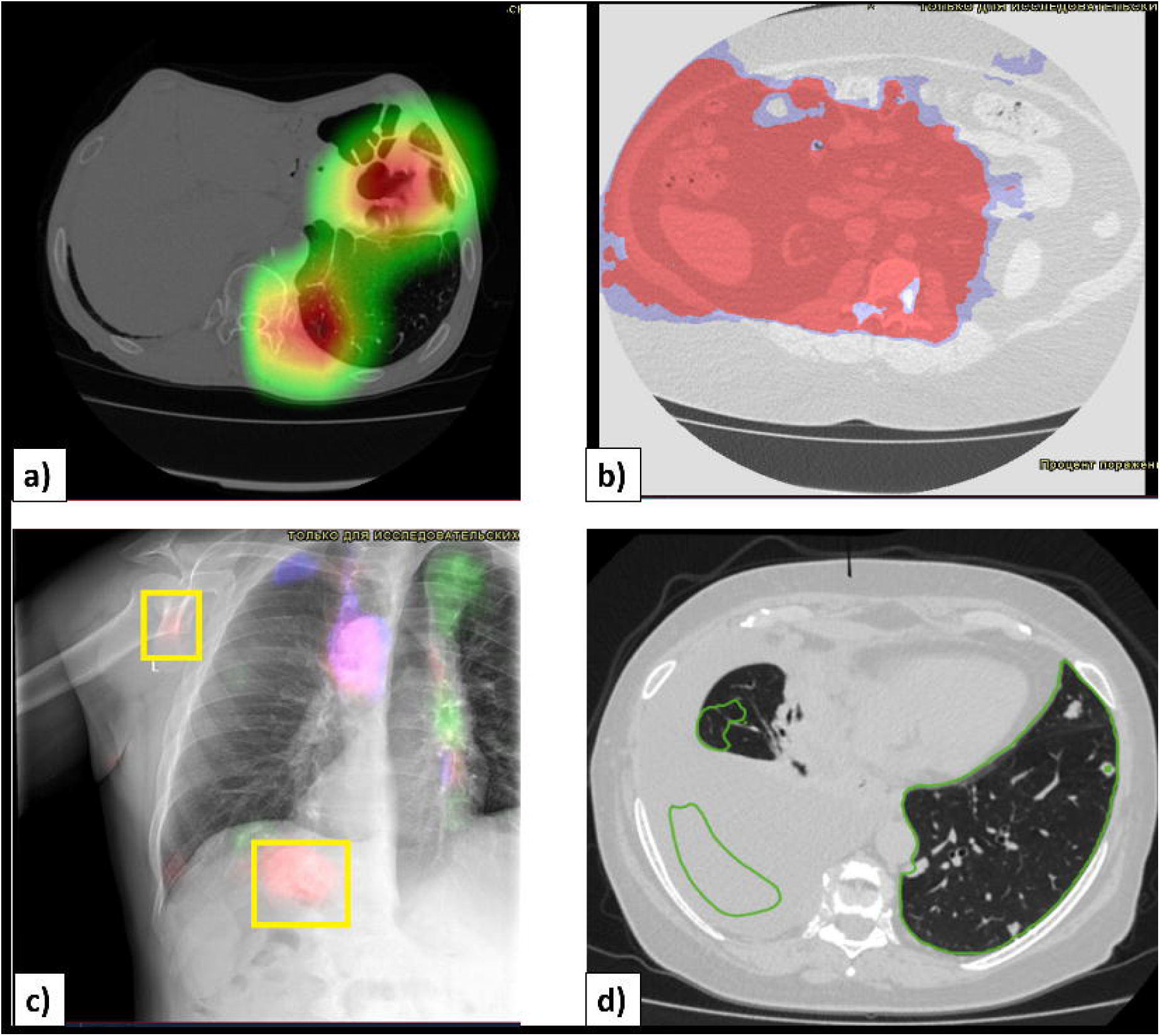
Examples of AI’s malperformance: a) and b) the AI marked the contents of bowels and abdominal organs, and incorrectly labeled them as pathological findings when processing a chest CT scan; c) when processing a chest X-ray, the AI marked the area outside a thorax, and incorrectly labeled the contents of bowels and a summation of the contours of humerus and coracoid process of the scapula; d) during a chest CT scan for detecting COVID-19 signs in the lungs, the AI labeled incorrectly right-sided hydrothorax – these changes are certainly pathological and located in the chest cavity, however, they are not related to the target findings.

**Figure 2.**
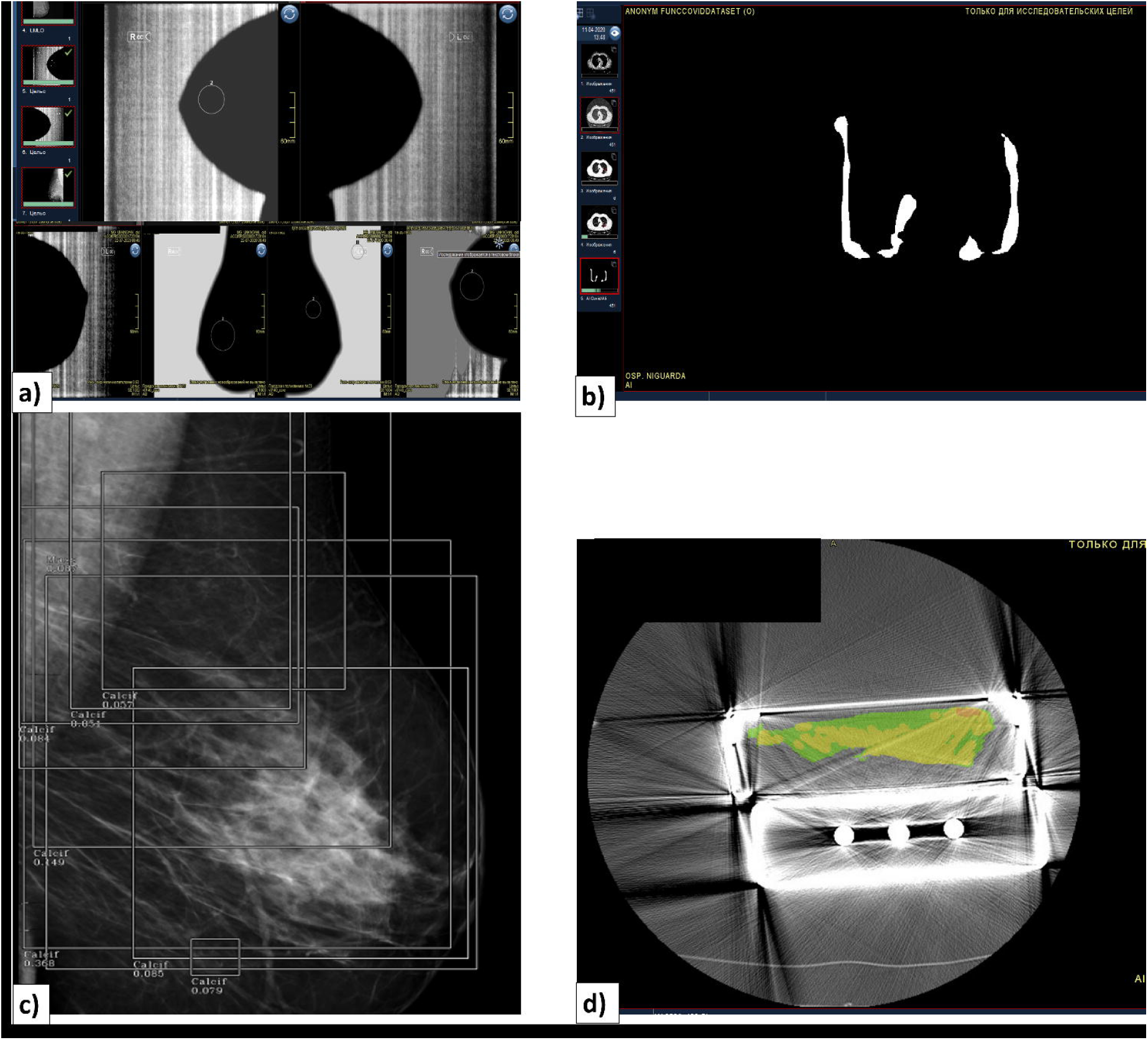
Examples of technincal defects that can be detected without a participation of the radiologist. Errors in the implementation of the AI functionality that make AI results unsuitable are demonstrated: a) and b) distortion of the original image when creating an additional series with labeling of pathological changes; c) incorrect labeling of findings and overlapping of labeling outlines; d) incorrect selection of images for the analysis – phantom’s images on CT scan.

**An AI-generated structure of the text report** (as part of DICOM SR) must contain:

– the text is presented in the national language. The experiment organizer’s staff-certified translators assisted the translation. Radiologists were involved in complicated cases.
– general information about AI – name, versioning, date and time of the beginning and completion of the analysis;
– a goal or clinical task solved by the AI;
– a brief user’s guide – for correct interpretation of radiological study images, as well as indication of a presence or absence of the additional series;
– conclusion and detailing of findings using generally accepted radiology terminology and systems, the same as used by radiologists in their reports.

*During the experiment, the report could be presented only on a separate tab of the URIS viewer. A proper visualization of the report on a tab in the URIS necessarily requires the appropriate desktop setting by the user before starting a work. However, individual developers provide an opportunity to receive a report in various text or graphic formats, online or by e-mail. In addition, a report might contain additional information and sections*.

#### 3. Calibration testing

assessed the expected diagnostic accuracy on a retrospective balanced local dataset of studies (100 studies, class balance – the ratio of studies with target changes and studies without changes 50/50). Within each diagnostic task, separate datasets were prepared. Each dataset was obtained from the URIS. The labeling was carried out by radiologists with 3 -10 years of experience (two per study with a third one in case of a disagreement) in double consensus and was based not only on presence/absence of radiological findings, but also on patient electronic medical records containing laboratory morphological verification, or based on the confirmation by the follow-up studies. Testing was carried out by data scientists. ROC-analysis was used to evaluate diagnostic accuracy metrics with the operation point determined by a maximum value of the Youden Index (J) for a local test dataset (10). Also, average processing times under the conditions simulating a real-life workflow were estimated. The processing time was measured between the time point a first image was sent to the AI up to the last processed image was received to “URIS-test”. The acceptable limit was set to 10 minutes.

**Figures 4a** shows the metrics (AUROC) of diagnostic accuracy for 40 AI that have undergone a calibration testing during the experiment. Only 15 out of 40 AI were approved for further clinical validation on prospective data, demonstrating a level of accuracy corresponding to AUROC above 0.81. Values below 0.6 characterized the AI as unsuitable for a further validation; AI with accuracy in the range of 0.61–0.8 required revision for readmission to the calibration testing(10).

**Figure 3.**
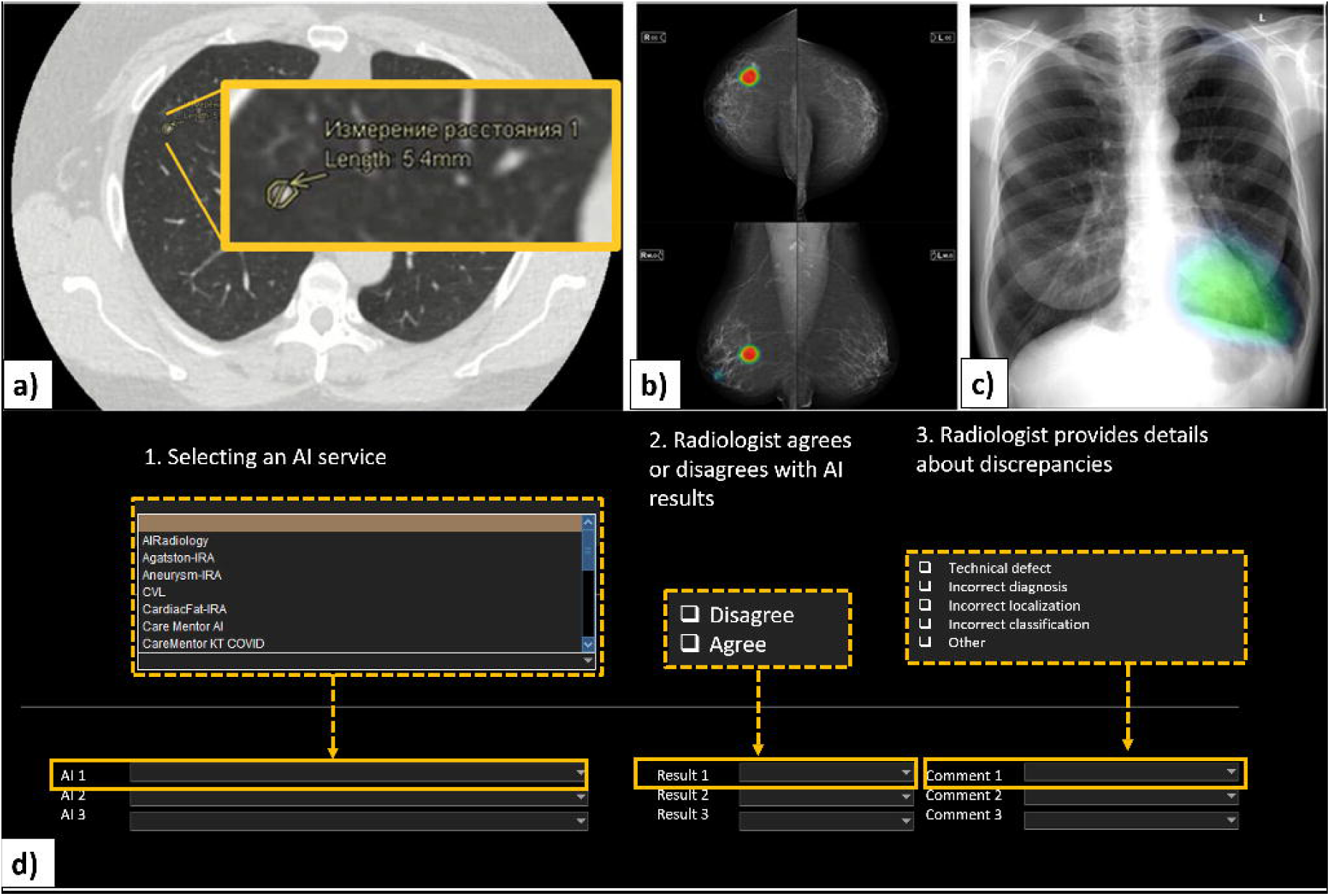
Examples of visualization of AI solutions integrated into RIS with labeling of pathological findings: a) chest CT scan; b) mammography exam; c) frontal chest X-ray; d) feedback questionnaire on the results of AI processing available to a radiologist.

**Figure 4.**
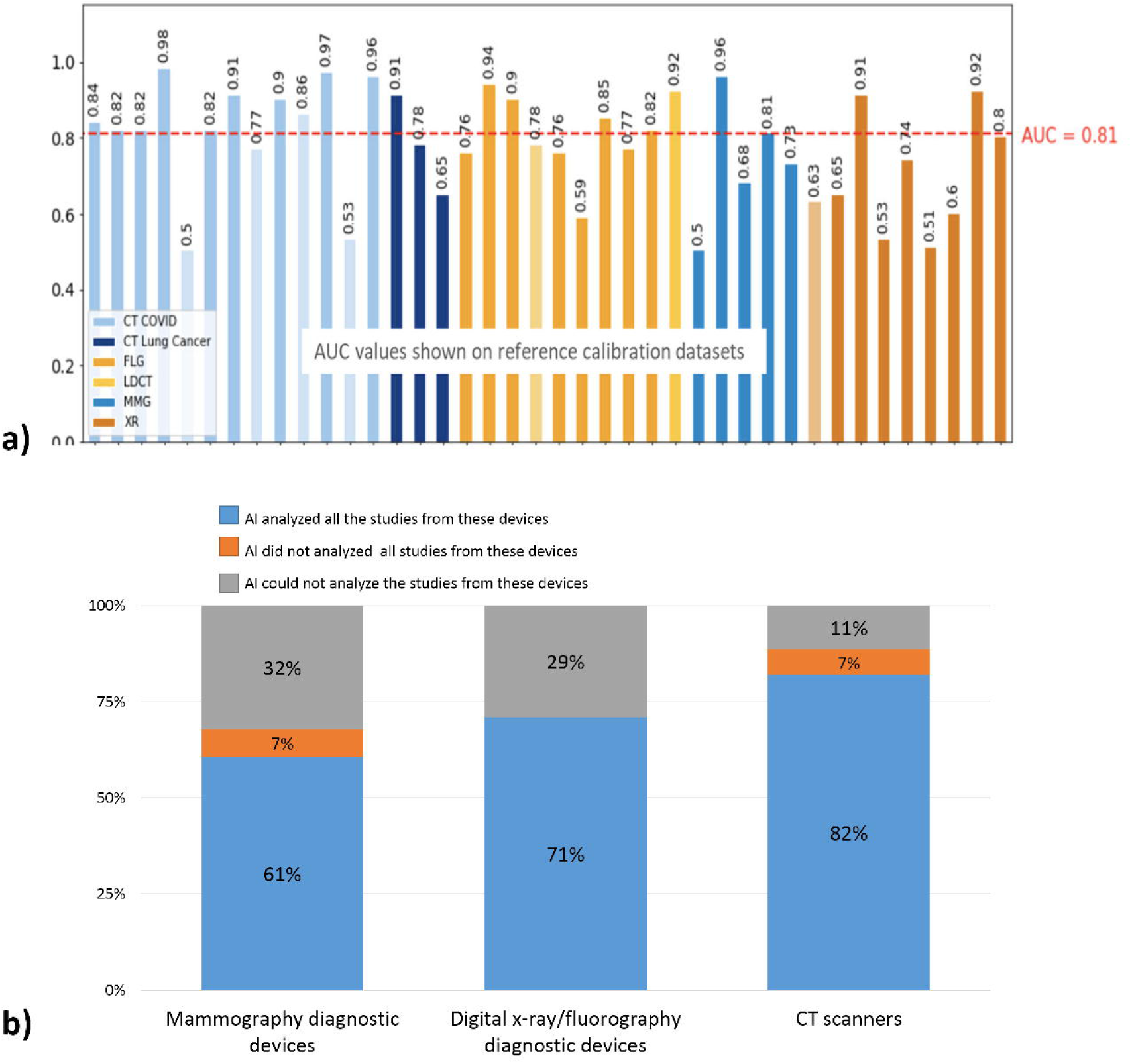
Diagnostic accuracy of AI solutions, expressed as the AUROC value according to the calibration testing data for CT, LDCT, X-ray, fluorography, and mammography modalities; Percentage of diagnostic devices from which studies could be processed, partially processed, or not processed by AI solutions.

#### 4. Technical testing and monitoring

When the AI is embedded into actual radiology workflow, TT and periodic monitoring of technological defects ensure stability and quality control during AI operation on real-time data.

TT was always carried out when an AI solution or its update received studies for a first time from the particular diagnostic device. TT was done based on 2 studies per device. A total of 20 TT procedures were performed covering 295 diagnostic devices (of which CT – 32.0%, X-ray/ FLG – 53.0%, MMG – 15.0%). 11% of CT, 29% of X-ray and 32% of mammography devices studies appeared to be incompatible with AI(**Fig. 4b**). 7% of mammography units and 7% of CT-scanners demonstrated variability in operating with the different AI models. These results indicate the necessity to include examples from all diagnostic devices instead of per model during the self-test stage.

Technical failures of AI models performance were evaluated automatically during monitoring. They were divided into groups: defects related to increase in the processing time and failures to analyze a study/deliver results to URIS (assessed by data scientists), and malperformance of AI (assessed by radiologists). Monitoring was carried out on a regular basis, its frequency varied from weekly to monthly. Identified defects were reported and sent to the AI providers. When comparing the number of detected defects, a decrease was noted with each subsequent monitoring **(Fig. 5)**. If a level of defects persisted, a decision was made to disconnect this AI from diagnostic devices.

**Figure 5.**
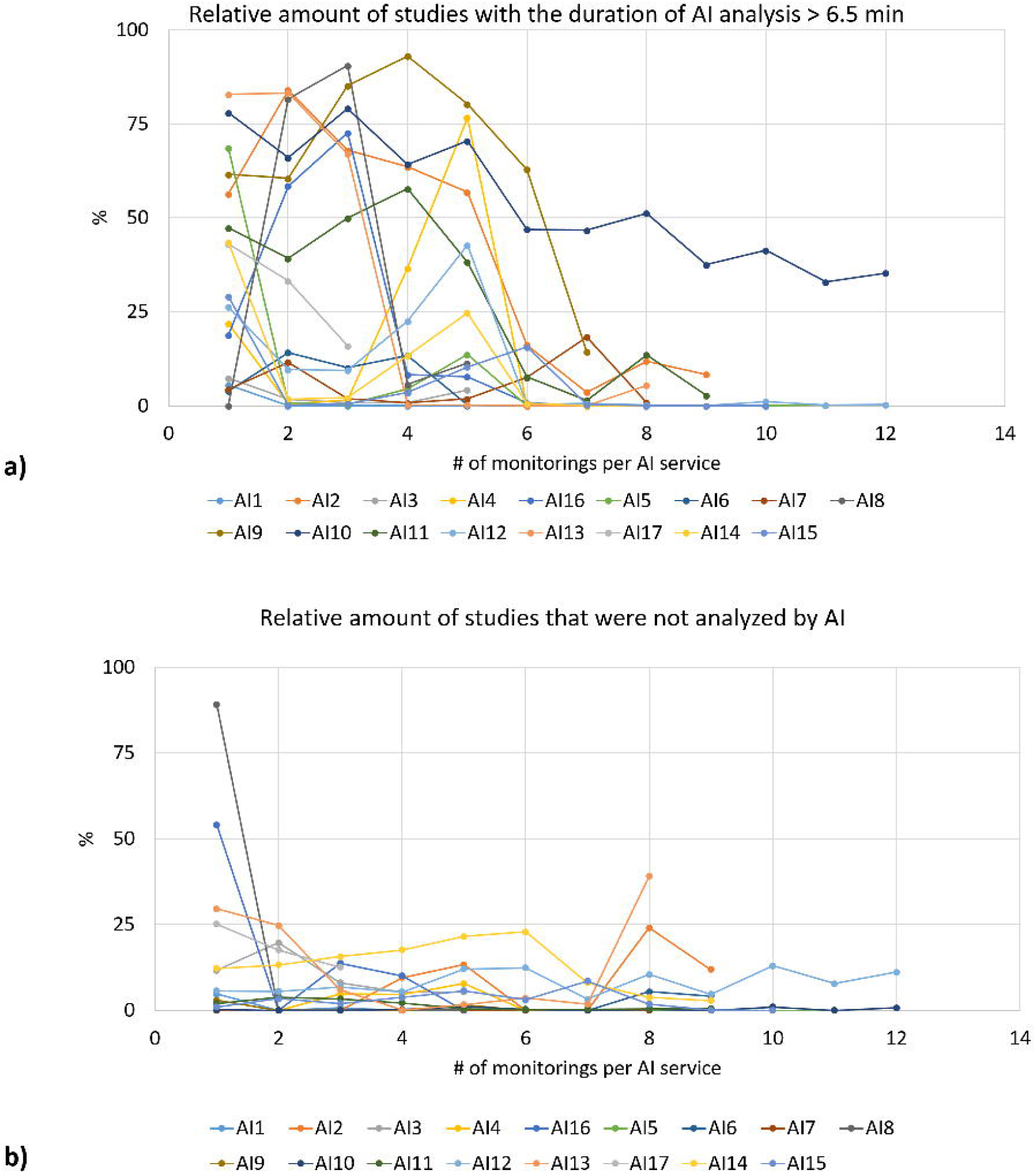
Positive dynamics of improving AI reliability during periodic monitoring of workflow and feedback for suppliers. a) A decrease in the studies taking more than 6.5 minutes for AI-processing is noted for most of the AI solutions. b) There is a decrease in the number of unanalyzed studies.

*The abundance of defects made it necessary to increase a monitoring frequency up to once a week and to notify the AI model provider about encountered malfunctioning more often. With the refinement of AI model, a number of defects and a frequency of monitoring decreased. Standard monitoring was conducted once a month, and if the number of defects exceeded 10%, the frequency of such analysis was reduced back to 1 week. This weekly monitoring continued until the relative number of defects dropped below 10%. Sudden increase in the number of defects could occur after the software changes*.

#### 5. Feedback from radiologists and an expert audit

To assess a practical value of the tested AI, a possibility to give feedback was provided for radiologists, followed by an expert audit of the selected AI results.

Feedback from radiologists was collected for the processed studies in real-time without interrupting the usual workflow by integrating a feedback questionnaire into the URIS (**Fig 3d**). Every radiologist could leave feedback on every result of the AI algorithm directly in the URIS (a special section with feedback fields was added). Radiologists could agree or disagree with AI. In case of disagreement, a reason should have been chosen: presence of technological defect, or a discrepancy in the localization or classification (diagnosis) of findings. Technological defects based on the results of feedback analysis were confirmed in the same way as during monitoring. Discrepancies between AI and radiologists were verified by the expert audit of a sample of studies. A certified radiologist with the experience of more than 5 years in a specific sub-specialization, filled out a similar form, agreeing or disagreeing with the results of the radiologist and the AI, indicating a reason for disagreement.

## Discussion

The proposed methodology was tested at the scale of a large city and demonstrated its value in the selection and implementation of high quality and relevant AI solutions into a practice by eliminating coherently 25 out of 40 AI solutions (**Fig. 6**). The amount of cases required radiologists’ assessment was minimal since the methodology allowed reducing a workload on the overworked radiologists. Radiologists had the greatest influence on development of a concept and requirements for AI. The methodology is scalable and can be adapted for different ways of AI deployment in medicine, various users, diagnostic equipment and use cases.

**Figure 6.**
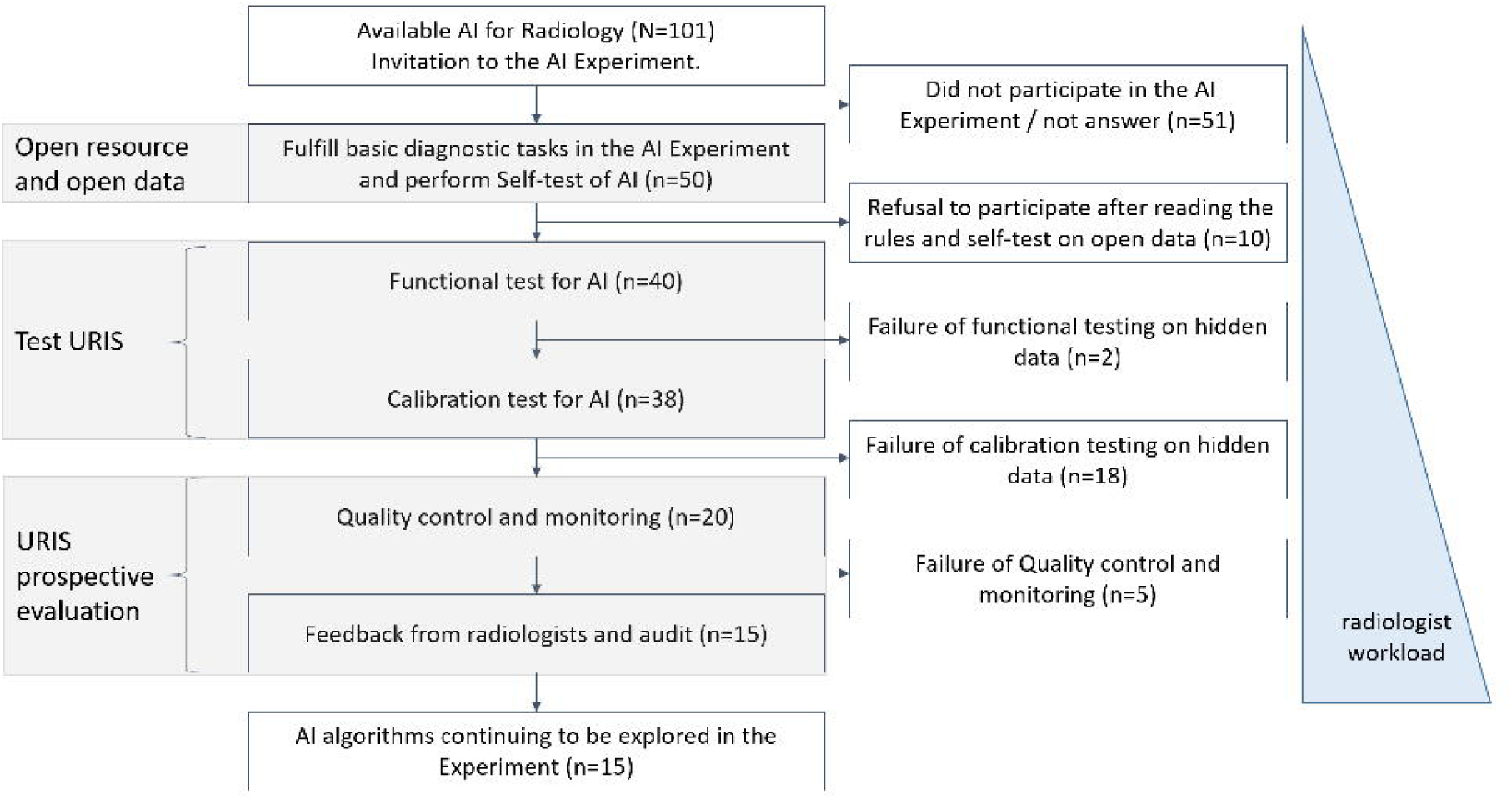
Scheme of a “sieve” for the selection process of AI solutions for clinical validation and implementation into practice by the proposed methodology.

A valuable feature of the developed methodology is a mandatory calibration on the local test dataset and subsequent validation on a real-time data accompanied by an expert audit. In the experiment, the high diagnostic accuracy metrics claimed by the developers was not confirmed for more than half of AI models. These solutions were excluded from the further validation, which allowed saving resources and making the integration safe for patients. It corresponds to the world experience. Most of the research on the use of AI in medicine did not contain a comparative analysis of the capabilities of AI and physicians, did not provide detailed information on the size and structure of samples for a comparative analysis(14); the majority of studies did not contain the results of external validation and AI application in practice; and in contrast to metrics claimed by the developers, less than 1% of the algorithms can be considered reliable according to the independent assessment(14). In 2018, it was noted that a quality of the external validation leaves much to be desired – it was carried out only in 6% of studies, among them none of the studies contained simultaneously a diagnostic cohort design, inclusion of multiple institutions, and for prospective data collection 99% of studies were built in accordance with proof-of-concept and feasibility study designs(15).

According to Gartner Hype Cycle, AI solutions for diagnostic radiology that do not meet the inflated users’ expectations inevitably dampens enthusiasm and creates obstacles to the further development and formation of new technologies(16). Developed and tested methodology contributes to the selection of the most relevant and high-quality AI. Its further improvement will help mitigate user dissatisfaction with the discrepancy between the AI’s functionality and expectations and could allow to painlessly overcome a decline of interest in AI technologies and lead to a speedy development of AI and its safe and stable performance in practice.

A stability and quality of AI results depend on the quality of incoming data. During the experiment, the stability of data transfer between diagnostic devices and AI was provided and supported by the URIS. For mammography and X-ray devices, a share of stably connected equipment was 61-71%, for СT-scanners – 82%. Preliminary quality control is possible either from the developers’ side or inside specially designed modules integrated with the URIS. The step-by-step implementation is aimed to detect and exclude AI solutions that do not meet the basic requirements, by a transition from the simplest and fastest tests on a small dataset to more labor-intensive ones that involve the experts of different specialties. Thus, only AI solutions meeting the basic requirements were admitted for a thorough analysis by data scientists. And only AIs with the acceptable diagnostic accuracy were admitted for an expert analysis by radiologists. The urgency of saving time of highly qualified specialists and their redistribution to more complex and precise diagnostic tasks in the implementation of large projects is beyond doubt. The ability to focus the specialist’s attention on complex tasks will lead to the care quality improvement in the primary health care through a reallocation of labor resources. Prospects for improving this methodology in the form of markup localization control, as well as quantification of AI performance. In long term, it becomes efficient for the end user – including the administration of the medical facility.

## Summary Statement

Practical implementation of radiological AI on large non-academic scale revealed critical issues for user-radiologists, IT-specialists and AI-developers perspectives that could be addressed via a structured methodology for selection and evaluation.

## Data Availability

All data produced in the present study are available upon reasonable request to the authors

## List of abbreviations

AI: artificial intelligence
ASPECTS: The Alberta stroke program early CT score
AUROC: area under receiver operating curve
CT: computed tomography
DICOM: digital imaging and communications in medicine
DICOM SR: digital imaging and communications in medicine structured report
FLG: fluorography
LDCT: low-dose CT
MMG: mammography
PACS: picture archiving computer system
QC: quality control
RIS: Radiological Information System
TT: technical testing
URIS: the Unified Radiological Information Service

## Notes

### Competing Interest Statement

The authors have declared no competing interest.

### Funding Statement

This study did not receive any funding

### Author Declarations

Independent Ethics Committee (IEC) of Moscow Regional Office (MRO) of the Russian Society of Radiologists and Radiographists (RSRR) gave ethical approval for this work as part of the Experiment on the use of innovative computer vision technologies for medical image analysis and subsequent applicability in the healthcare system of Moscow. Details of the IEC: Approval Number No.2 (1-II-2020) of the meeting of IEC of RSRR MRO dated February 20, 2020. Address: 28 Srednyaya Kalitnikovskaya St., bld. 1, Moscow. Email: ethics{at}npcmr.ru Present: O.A. Agafonova, E.G. Bahteeva, A.S. Laipan, I.S. Komolov, O.A. Mokienko, I.I. Nadelyaeva, L.A. Nizovtsova, O.V. Omelyanskaya, A.V. Petraikin. Topics: Expert review of the documents for the upcoming study titled the "Experiment on the use of innovative technologies in the field of computer vision for the analysis of medical images and further use in the healthcare system of Moscow." Documents submitted for review: 1. The abstract of the study titled "Experiment on the use of innovative technologies in the field of computer vision for the analysis of medical images and further use in the healthcare system of Moscow" 2. A cover letter to the attention of the Chairman of IEC of RSRR MRO concerning the erhical expert review of the study titled the "Experiment on the use of innovative technologies in the field of computer vision for the analysis of medical images and further use in the healthcare system of Moscow" 3. CV of the Principal Investigator 4. A patient information and informed consent form 5. A project overview Resolution: To approve the upcoming study titled the "Experiment on the use of innovative technologies in the field of computer vision for the analysis of medical images and further use in the healthcare system of Moscow." Due to the conflict of interest O.A. Agafonova, the co-implementer of the project, did not take part in the vote. All necessary patient/participant consent has been obtained and the appropriate institutional forms have been archived.

